# Contact patterns of UK home delivery drivers and their use of protective measures during the COVID-19 pandemic: a cross-sectional study

**DOI:** 10.1101/2022.09.09.22279754

**Authors:** Jessica RE Bridgen, Hua Wei, Carl Whitfield, Yang Han, Ian Hall, Chris P Jewell, Martie JA van Tongeren, Jonathan M Read

## Abstract

**Objectives:** To quantify contact patterns of UK home delivery drivers and identify protective measures adopted during the pandemic.

**Methods:** We conducted a cross-sectional online survey to measure the interactions of 170 UK delivery drivers during a working shift between 7 December 2020 and 31 March 2021.

**Results:** Delivery drivers had a mean number of 71.6 (95% Confidence Interval (CI) 61.0 to 84.1) customer contacts per shift and 15.0 (95%CI 11.19 to 19.20) depot contacts per shift. Maintaining physical distancing with customers was more common than at delivery depots. Prolonged contact (more than 5 minutes) with customers was reported by 5.4% of drivers on their last shift. We found 3.0% of drivers had tested positive for SARS-CoV-2 since the start of the pandemic and 16.8% of drivers had self-isolated due to a suspected or confirmed case of COVID-19. Additionally, 5.3% (95%CI 2.3% to 10.2%) of participants reported having worked whilst ill with COVID-19 symptoms, or with a member of their household having a suspected or confirmed case of COVID-19.

**Conclusion:** Delivery drivers had a large number of face-to-face customer and depot contacts per shift compared to other working adults during this time. However, transmission risk may be curtailed as contact with customers was of short duration. Most drivers were unable to maintain physical distance with customers and at depots at all times. Usage of protective items such as face masks and hand sanitizer was widespread.

## INTRODUCTION

The delivery sector has been central in ensuring that services and supplies have remained available throughout the COVID-19 pandemic in the UK. There has been an unprecedented demand for home deliveries during the pandemic, rising sharply with the implementation of nationwide ‘stay at home’ orders.[1] The UK government classed delivery drivers as *key workers*, defined as workers critical to the COVID-19 response.[2] Shielding guidance for clinically extremely vulnerable individuals in January 2021 advised the use of food and prescription delivery services to minimise the need to leave home.[3] As non-essential businesses were forced to close for extended periods of time in 2020 and 2021, many businesses relied on online sales to generate income.[4]

Transmission of SARS-CoV-2, the virus that causes COVID-19, primarily occurs through airborne routes, however indirect transmission can take place through contaminated surfaces.[5,6] High contact occupations are thought to be associated with an increased likelihood of employees being exposed to SARS-CoV-2 and developing clusters of cases in the workplace.[7,8] Reducing the number of social contacts, increasing ventilation and frequent hand washing were advised methods to reduce workplace risk of exposure to SARS-CoV-2.[9] Delivery drivers interact regularly with other employees at depots (or collection hubs) and with a large number of customers. To mitigate against infection, contact-free deliveries became widely available to minimise contact and reduce transmission risk between delivery drivers and customers.[10] However, several studies have found that transport workers were at a higher risk of severe COVID-19 when compared with non-essential workers.[11,12]

We conducted a cross-sectional online survey between 7 December 2020 and 31 March 2021 to quantify behaviours thought to be associated with transmission risk of SARS-CoV-2. The study period coincides with the peak and gradual decline of the second wave of COVID-19 in the UK, following the emergence of the alpha variant. [13,14] The UK also entered a period of lockdown during this time, where ‘stay at home’ orders were in place, and non-essential businesses were closed to reduce transmission.[15] We aimed to quantify the contact patterns of delivery drivers within their depot and with their customers, identifying the types of contact they were making.

## METHODS

### Survey methodology

An anonymous online survey (the ‘CoCoNet: Home Delivery Driver survey’) was used for data collection. The survey design was adapted from a previous population-wide study.[16] Study participants had to meet the following inclusion criteria: a resident in the UK at the time of the survey, working as a home delivery driver and aged 18 or over. The survey was promoted through university press releases, engagement with delivery driver groups on social media (LinkedIn and Facebook) and targeted Facebook advertisements.

Survey responses received between 9 December 2020 and 31 March 2021 were included in the analysis. Partial survey responses were analysed for all questions that had been displayed to the participant.

Age, sex, ethnicity, household size and other demographic information was collected from participants. Employment information was requested, including employment type, working hours, types of items typically delivered and sick pay eligibility. The survey included questions pertaining directly to COVID-19, such as whether participants had tested positive for SARS-CoV-2, if they had to self-isolate due to suspected or confirmed SARS-CoV-2 infection and if they had worked whilst being ill with COVID-19 symptoms. Participants were asked to recall specific details from their last shift working as a delivery driver, including the number of customers they met face-to-face, the number of individuals they had a face-to-face conversation with at their depot and their use of personal protective equipment (PPE). The survey questions can be found in Supplementary Material.

### Primary and secondary outcome measurements

Our primary outcome measurement was the number of contacts delivery drivers have per shift. This was stratified into contact with customers and contact with individuals (employees or customers) at a delivery depot. A contact was defined as someone whom the participant had a face-to-face conversation with. Secondary outcome measurements were: the number of deliveries per shift; the type of contact drivers were having with customers; ability to maintain physical distance; use of protective items; COVID-related presenteeism; the frequency of self-isolation and COVID-19 infection.

### Data analysis

Study representativeness was assessed by comparing participant demographics with the Labour Force Survey (LFS) estimates for delivery drivers and couriers. Quarterly labour force surveys for the time period November 2020 to January 2022 were aggregated for comparison, due to the relatively small sample size of delivery drivers and couriers included in each individual survey.[17–22]

To identify occupational and personal characteristics associated with participants’ interactions with customers, we fitted a negative binomial regression model to the number of customer contacts per shift. Explanatory variables included in the model were: age, sex, employment type, furthest distance travelled from the collection point to a delivery, weekly working hours and the type of items delivered. The model was assessed for multicollinearity by calculating the variance inflation factor for each independent variable.

### Ethics statement

This study was approved by the Faculty of Health and Medicine Ethics Committee at Lancaster University (reference FHMREC20040). Participation in the study was voluntary and all participants provided consent before proceeding with the survey.

## RESULTS

### Participant demographics

We received 170 survey responses between 9 December 2020 and 31 March 2021 which met our inclusion criteria. Male participants accounted for 75.3% (128/170) of the sample, our survey over sampled females when compared to the aggregated LFS; Table 1. The majority of participants were aged 40 to 59 (56.4%, 96/170). Participants predominantly resided in England (81.8%, 139/170), with 1.8% (3/170) of participants residing in Northern Ireland, 10.6% (18/170) residing in Scotland and 5.9% (10/170) residing in Wales. We found that our sample was representative by ethnicity and nation when compared to the population estimates of delivery drivers as reported by the aggregated LFS; Table 1.

**Table 1:** Participant demographics and aggregated labour force survey estimates for ‘delivery drivers and couriers’. N is the number of participants who provided a response to the question.

|  | Number of participants<br>(percentage, 95% binomial confidence interval) | Aggregated<br>Quarterly Labour Force Survey<br>estimates for<br>delivery drivers<br>and couriers* [17–22]<br>November 2020 - January 2022<br>Number of participants<br>(percentage) |
| --- | --- | --- |
| Age group | N = 170<sup>†</sup> | N = 1785 |
| <18 | - ** | 5 (0.3%) |
| 18-29 | 27 (15.9%, 10.74%-22.26%) | 230 (12.9%) |
| 30-39 | 35 (20.6%, 14.78%-27.45%) | 245 (13.7%) |
| 40-49 | 46 (27.1%, 20.54%-34.39%) | 338 (18.9%) |
| 50-59 | 50 (29.4%, 22.68%-36.87%) | 530 (29.7%) |
| 60-69 | 12 (7.1%, 3.70%-12.01%) | 384 (21.5%) |
| 70+ | 0 (0.0%, 0.00%-2.15%) | 53 (3.0%) |
| Sex | N = 170<sup>†</sup> | N = 1785 |
| Female | 40 (23.5%, 17.37%-30.63%) | 181 (10.1%) |
| Male | 128 (75.3%, 68.11%-81.58%) | 1604 (89.9%) |
| Prefer not to say | 2 (1.2%, 0.14%-4.19%) | - |
| Ethnicity | N = 170 | N = 1785 |
| White | 156 (91.8%, 86.57%-95.42%) | 1648 (92.3%) |
| Mixed/Multiple ethnic groups | 5 (2.9%, 0.96%-6.73%) | 9 (0.5%) |
| Asian/Asian British | 5 (2.9%, 0.96%-6.73%) | 88 (4.9%) |
| Black/African/Caribbean/Black British | 1 (0.6%, 0.01%-3.23%) | 26 (1.5%) |
| Other ethnic groups | 0 (0.0%, 0.00%-2.15%) | 14 (0.8%) |
| Prefer not to say | 3 (1.8%, 0.37%-5.07%) | - |
| No response | 0.00 (0%, 0.00%-2.15%) | 0 (0.0%) |
| Nation | N = 170<sup>†</sup> | N = 1785 |
| England | 139 (81.8%, 75.13%-87.26%) | 1491 (83.5%) |
| Northern Ireland | 3 (1.8%, 0.37%-5.07%) | 79 (4.4%) |
| Scotland | 18 (10.6%, 6.40%-16.22%) | 114 (6.4%) |
| Wales | 10 (5.9%, 2.86%-10.55%) | 101 (5.7%) |

\* Main occupation of participant recorded as 'delivery drivers and couriers'
<sup>†</sup> Question required a response from the participant
\*\* Age group did not meet study inclusion criteria

### Employment situation

The majority of participants reported their employment situation to be either self-employed and completely independent (54.3%, 95% confidence interval (CI) 46.3% to 62.2%) or employed full time by one company (28.4%, 95%CI 21.6% to 36.0%); see supplementary material Table 1. Over half of delivery drivers (53.1%, 95%CI 45.1% to 61.0%) reported their weekly working hours to be between 31 and 50 hours, and 22.2% (95%CI 16.1% to 29.4%) of delivery drivers reported working more than 50 hours a week. The majority of participants reported their most recent working shift to be during the week of completing the survey (92.6%, 95%CI 87.4% to 96.1%). A small proportion of participants reported their most recent working shift to be more than a month before completing the survey (2.5%, 95%CI 0.7% to 6.2%). Most participants (68.2%, 95%CI 60.1% to 75.5%) reported that they did not receive statutory sick leave pay whilst working as delivery drivers.

### Workplace interactions

The mean number of customer contacts was 71.6 (95%CI 61.0 to 84.1) per shift. We found 95.2% (95%CI 90.4% to 98.1%) of participants had brief face-to-face interactions (less than 5 minutes) with customers on their last shift, 5.4% (95%CI 2.4% to 10.4%) of participants had prolonged face-to-face interactions (more than 5 minutes) with customers and 8.2% (95%CI 4.3% to 13.8%) had entered a customer’s property. We found that 61.9% (95%CI 53.5% to 69.8%) of participants were able to maintain physical distance with customers at all times during their last shift. A small proportion of participants (2.7%, 95%CI 0.7% to 6.8%) reported that they could not maintain physical distance at all on their last shift.

The mean number of contacts per shift in depot (where drivers collected items for delivery) was 27.9 (95%CI 12.2 to 55.8). This was reduced to 15.0 (95%CI 11.19 to 19.20) contacts per shift after excluding a single individual who reported making an exceptionally large number of contacts. We found that 42.4% (95%CI 34.2% to 50.9%) of participants reported that they were able to maintain physical distance from contacts at the depot at all times during their last shift. Whereas 8.3% (95%CI 4.4% to 14.1%) of participants were unable to maintain physical distance at all on their previous shift.

We found 10.5% (95%CI 6.1% to 16.4%) of participants shared a vehicle with a colleague during their last working week, of which, 56.2% (95%CI 29.9% to 80.2%) reported sharing the vehicle with the same colleague throughout the week.

The number of contacts made per shift, including both customer and depot interactions, was positively correlated with the number of deliveries made per shift; Figure 1A. Participants who reported typically delivering only large items had the greatest number of customer and depot contacts per shift, making on average more customer contacts than deliveries per shift; Figure 1B. While most drivers delivering large items only reported a one-to-one ratio of customer contacts and deliveries, one individual reported four times the number of customer contacts than deliveries.

**Figure 1:**
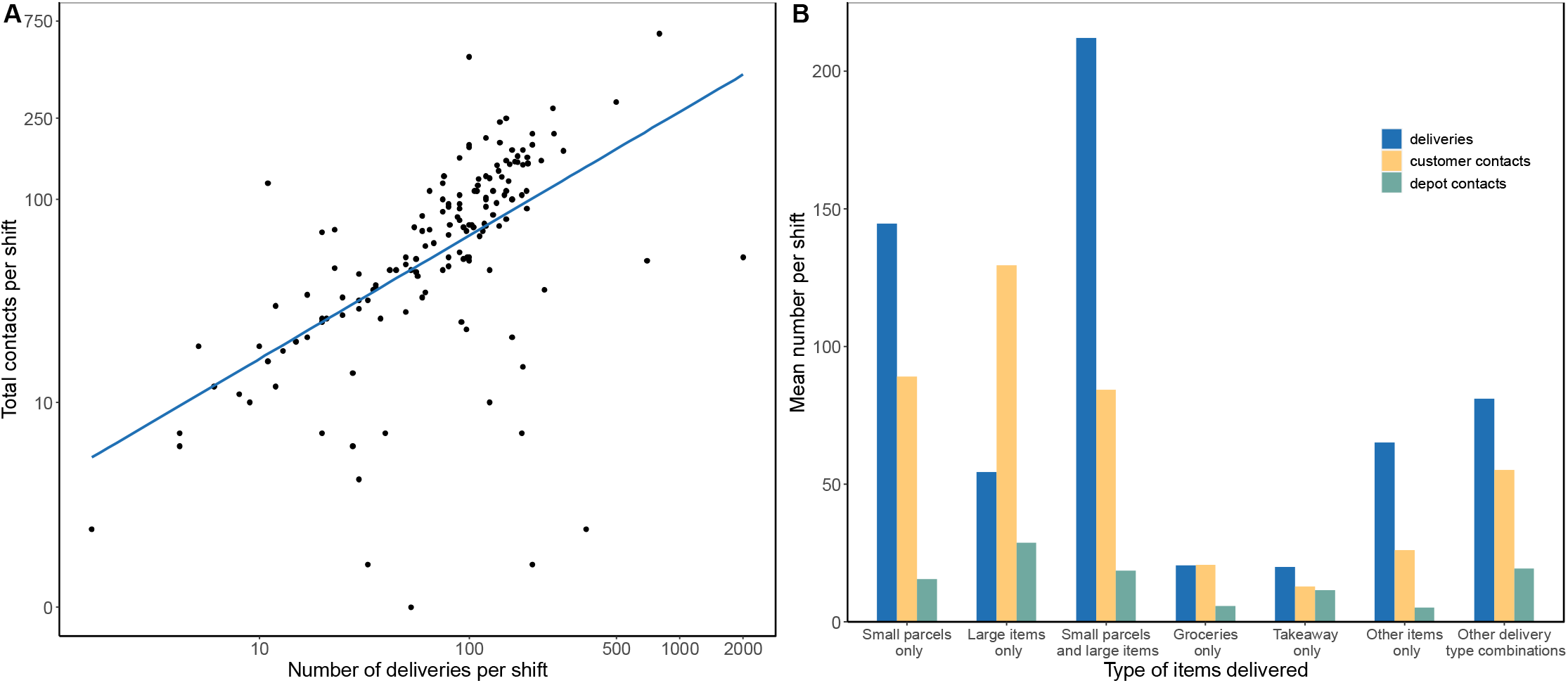
A) Number of total contacts and deliveries made per shift. Note, x-axis and y-axis on log-scale. B) Mean number of deliveries and contacts per shift by delivery type.

### Frequency and type of deliveries

We found that the mean number of deliveries per shift was 121.8 (95%CI 97.9 to 152.3). Approximately half of participants (51.0%, 95%CI 42.8% to 59.1%) reported that the furthest distance they travelled from a collection point to a delivery address during their last working week was under 20 miles. The majority of delivery drivers surveyed (52.5%, 95%CI 44.5% to 60.4%) reported that they typically delivered small parcels (including letters and mail). We found that drivers delivering small parcels and large items had the highest mean number of deliveries per shift, whilst takeaway and grocery delivery drivers had the lowest; Figure 1B.

### Predictors of customer contacts

A negative binomial model was fitted to the number of face-to-face customer interactions per shift. The variance inflation factor was less than five for all model coefficients indicating multicollinearity was not present. Participants aged 18 to 29 (adjusted incidence rate ratio (aIRR) 1.65, 95%CI 1.07 to 2.06) and aged 40 to 49 (aIRR 1.64, 95%CI 1.15 to 2.34) had a higher number of customer contacts per shift than those aged 50 to 59; Figure 2, supplementary material Table 2. We found that delivery drivers who were employed by one company full time had a lower number of customer contacts per shift than those self-employed and independent (aIRR 0.66, 95% CI 0.47 to 0.94). Participants who usually deliver only groceries (aIRR 0.34, 95%CI 0.18 to 0.64), deliver only takeaways (aIRR 0.19, 95%CI 0.11 to 0.37), deliver other unlisted items (aIRR 0.43, 95%CI 0.24 to 0.80) and those who deliver other combinations of items listed (aIRR 0.58, 95%CI 0.38 to 0.93) had fewer customer contacts than those that usually delivery only small parcels.

**Figure 2:**
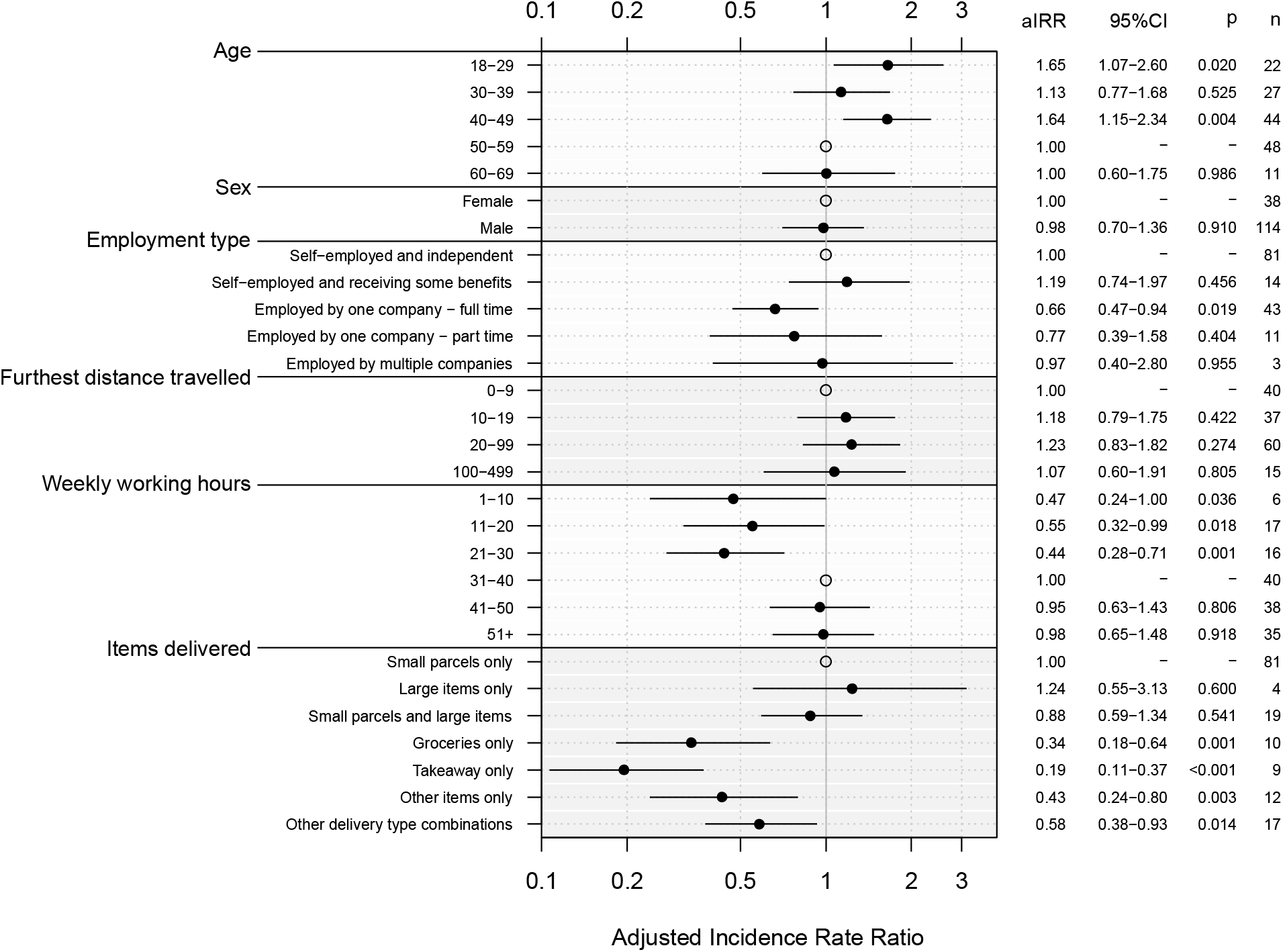
Adjusted incidence rate ratios for mean number of customer contacts reported for selected variables. Open circles represent the reference group for each variable.

### COVID-19 infection, self-isolation and presenteeism

We found that 3.0% (95%CI 1.0% to 6.9%) of delivery drivers surveyed reported that they had tested positive for COVID-19. Moreover, 16.8% (95%CI 11.4% to 23.3%) of delivery drivers had self-isolated since the start of the pandemic due to a suspected or confirmed case of COVID-19. Approximately one in twenty drivers (5.3%, 95%CI 2.3% to 10.2%) reported that they have continued to work whilst being ill with COVID-19 symptoms or with a member of their household having a suspected or confirmed case of COVID-19. In this situation, financial reasons were most often cited as a reason for continuing to work (85.7%, 95%CI 42.1% to 99.6%).

### Protective measures

We found that 68.3% (95%CI 60.0% to 75.7%) of participants were provided with some PPE items by their employers or contracting companies. However, less than half of participants (48.3%, 95%CI 39.9% to 56.7%) felt that the PPE provided effective protection. Facemasks (82.4%, 95%CI 75.4% to 88.0%) and hand sanitiser (83.7%, 95%CI 76.8% to 89.1%) were the protective items most commonly used by delivery drivers on their last shift. Participants who shared a vehicle during their last working week most often reported using hand sanitizer to prevent infection when sharing a vehicle (81.2%, 95%CI 54.3% to 96.0%), with 50.0% (95%CI 24.7% to 75.3%) of participants reporting wearing a facemask and 50.0% (95%CI 24.7% to 75.3%) reporting keeping a window open.

## DISCUSSION

We found that delivery drivers made a large number of contacts per shift both at their depot (15.0 per shift) and with customers (71.6 per shift). In comparison, Jarvis *et al*. found that the mean number of contacts among the general population attending their workplace between January 2021 and March 2021 was between 3 and 10 contacts per day; this included contacts made outside of the workplace [23]. This suggests delivery drivers have a very large number of contacts compared to the general workforce at this time, which may lead to a higher risk of SARS-CoV-2 infection. The importance of contact duration in respiratory virus transmission has been widely documented.[24–28] Face-to-face interactions between delivery drivers and customers are likely to take place outside and to be very short in duration, with only 5.4% of drivers reporting any prolonged contact (more than 5 minutes) with customers during their last working shift. Therefore, whilst delivery drivers have a large number of contacts this may pose only a small risk in terms of exposure and transmission of SARS-CoV-2. Similarly, sharing a vehicle with a colleague for deliveries may be a type of high-risk contact. Nevertheless, as we found most work-related vehicle sharing was fixed-pairing (pair that share a vehicle is fixed) the risk of multiple transmission events is likely to be largely reduced. The duration of contacts made at the depot was not recorded.

The use of protective measures in the workplace was common. Most delivery drivers reported being able to maintain physical distance with customers and at the depot at least some of the time during their last shift, however, most drivers reported not being able to maintain distance at all times particularly when at the depot. Facemasks and hand sanitiser were commonly used by drivers during their shift. While facemasks offer varying levels of protection to the wearer they help to prevent transmission from an infected individual to others.[29] The majority of drivers received some items of protective items from their employers, however, less than half of drivers felt that this provided effective protection.

By 7 December 2020, there had been 1,770,619 confirmed cases of COVID-19 in the UK, accounting for approximately 2.6% of the UK population.[13,30] We found 3.0% of delivery drivers surveyed had tested positive for SARS-CoV-2 since the start of the pandemic and over a sixth of delivery drivers reported having to self-isolate due to a suspected or confirmed case of SARS-CoV-2. A small proportion of delivery drivers reported working with symptoms of COVID-19 or while a member of their household had a confirmed or suspected SARS-CoV-2 infection. The majority of drivers reported continuing to work in this situation due to financial reasons, this may be associated with statutory sick pay being unavailable for most drivers. Lack of access to paid sick leave is one of the main risk factors for respiratory infectious disease-related presenteeism.[31] Providing access to paid sick leave may therefore help to improve adherence to public health measures such as self-isolation.

Participants self-reported how many face-to-face interactions they had with customers and at the depot on their last working shift as a delivery driver. For most participants their last working shift was during the same week as completing the survey, but a small proportion (approximately 2.5%) were recalling from a shift over a month ago. There is some risk of uncertainty in recall particularly with the small proportion of participants recalling from a less recent shift. This study may suffer from recruitment bias, the survey was conducted online only without a strict recruitment process. Mean number of contacts were calculated per shift and therefore cannot be directly compared with other contact studies which calculate the number of contacts per hour or per day. Questions pertaining to SARS-CoV-2 infection and self-isolation referred to the time period from the start of the pandemic to completing the survey. We did not collect data on how long participants had been working as delivery drivers, therefore estimations of the prevalence of SARS-CoV-2 infection and self-isolation amongst participants may not be an accurate representation of all delivery drivers.

## Supporting information

STROBE checklist

## Data Availability

Data are available in a public, open access repository. The survey data are available online from Lancaster University research directory. License: Creative Commons Attribution licence (CC BY).

https://doi.org/10.17635/lancaster/researchdata/553

## Acknowledgements

We would like to thank the participants of the study for their time and information.

## Contributions

All authors conceived and designed the study. JREB conducted the analysis and wrote the first draft of the manuscript. All authors edited the manuscript.

## Data Availability Statement

Data are available in a public, open access repository. The survey data are available from Lancaster University’s research directory at: https://doi.org/10.17635/lancaster/researchdata/553. License: Creative Commons Attribution licence (CC BY).

## References

1 Office for National Statistics. Impact of the coronavirus (COVID-19) pandemic on retail sales in 2020. 2021.https://www.ons.gov.uk/economy/grossdomesticproductgdp/articles/impactofthecoronaviruscovid19pandemiconretailsalesin2020/2021-01-28 (accessed Sep 2021).

2 GOV UK. Children of critical workers and vulnerable children who can access schools or educational settings. Gov.uk. https://www.gov.uk/government/publications/coronavirus-covid-19-maintaining-educational-provision/guidance-for-schools-colleges-and-local-authorities-on-maintaining-educational-provision (accessed Jan 2022).

3 Office for National Statistics. Coronavirus and clinically extremely vulnerable people in England - Office for National Statistics. 2021.https://www.ons.gov.uk/peoplepopulationandcommunity/healthandsocialcare/conditionsanddiseases/bulletins/coronavirusandclinicallyextremelyvulnerablepeopleinengland/11octoberto16october2021 (accessed Jan 2022).

4 Coronavirus: how people and businesses have adapted to lockdowns. 2021.https://www.ons.gov.uk/economy/economicoutputandproductivity/output/articles/coronavirushowpeopleandbusinesseshaveadaptedtolockdowns/2021-03-19 (accessed 27 Jan 2022).

5 World Health Organization. Transmission of SARS-CoV-2: implications for infection prevention precautions. 2020.https://www.who.int/news-room/commentaries/detail/transmission-of-sars-cov-2-implications-for-infection-prevention-precautions (accessed Aug 2021).

6 GOV UK. COVID-19: epidemiology, virology and clinical features. Gov.uk. 2022.https://www.gov.uk/government/publications/wuhan-novel-coronavirus-background-information/wuhan-novel-coronavirus-epidemiology-virology-and-clinical-features (accessed 2022).

7 Office for National Statistics. Which occupations have the highest potential exposure to the coronavirus (COVID-19)? 2020.https://www.ons.gov.uk/employmentandlabourmarket/peopleinwork/employmentandemployeetypes/articles/whichoccupationshavethehighestpotentialexposuretothecoronaviruscovid19/2020-05-11 (accessed Aug 2020).

8 European Centre for Disease Prevention and Control. COVID-19 clusters and outbreaks in occupational settings in the EU/EEA and the UK. 2020.https://www.ecdc.europa.eu/en/publications-data/covid-19-clusters-and-outbreaks-occupational-settings-eueea-and-uk (accessed 2021).

9 Health and Safety Executive. Working safely during the coronavirus (COVID-19) pandemic. https://www.hse.gov.uk/coronavirus/working-safely/index.htm (accessed Jan 2022).

10 Wei H, Daniels S, Whitfield CA, et al. Agility and Sustainability: A Qualitative Evaluation of COVID-19 Non-pharmaceutical Interventions in the UK Logistics Sector. Front Public Health 2022;10:864506.

11 Mutambudzi M, Niedwiedz C, Macdonald EB, et al. Occupation and risk of severe COVID-19: prospective cohort study of 120 075 UK Biobank participants. Occup Environ Med Published Online First: 9 December 2020. doi:10.1136/oemed-2020-106731

12 Nafilyan V, Pawelek P, Ayoubkhani D, et al. Occupation and COVID-19 mortality in England: a national linked data study of 14.3 million adults. Occup Environ Med Published Online First: 27 December 2021. doi:10.1136/oemed-2021-107818

13 Public Health England. Coronavirus (COVID-19) in the UK. 2020.https://coronavirus.data.gov.uk (accessed Aug 2020).

14 Public Health England. Investigation of SARS-CoV-2 variants of concern in England: technical briefing 6. 2021.

15 GOV UK. Prime Minister’s address to the nation: 4 January 2021. Gov.uk. 2021.https://www.gov.uk/government/speeches/prime-ministers-address-to-the-nation-4-january-2021 (accessed Jan 2022).

16 Bridgen JRE, Jewell CP, Read JM. Social mixing patterns in the UK following the relaxation of COVID-19 pandemic restrictions: a cross-sectional online survey. MedRxiv. 2021. doi:10.1101/2021.10.22.21265371

17 Northern Ireland Statistics and Research Agency (NISRA), Office for National Statistics, Social Survey Division. Quarterly Labour Force Survey, November, 2020 - January, 2021. UK Data Service. 2021. doi:10.5255/UKDA-SN-8788-2

18 Office for National Statistics, Social Survey Division, Northern Ireland Statistics and Research Agency, Central Survey Unit. Quarterly Labour Force Survey, February - April, 2021. UK Data Service. 2021. doi:10.5255/UKDA-SN-8813-2

19 Office for National Statistics, Northern Ireland Statistics and Research Agency. Quarterly Labour Force Survey, May - July, 2021. UK Data Service. 2021. doi:10.5255/UKDA-SN-8845-1

20 Office for National Statistics, Northern Ireland Statistics and Research Agency. Quarterly Labour Force Survey, July - September, 2021. UK Data ServiceUK Data Service. 2021. doi:10.5255/UKDA-SN-8872-1

21 Office for National Statistics, Northern Ireland Statistics and Research Agency. Quarterly Labour Force Survey, September - November 2021. UK Data Service. 2022. doi:10.5255/UKDA-SN-8901-1

22 Northern Ireland Statistics and Research Agency (NISRA), Office for National Statistics, Social Survey Division. Quarterly Labour Force Survey, November, 2021 - January, 2022. UK Data Service. 2022. doi: DOI: 10.5255/UKDA-SN-8927-2

23 Jarvis C, Edmunds J, CMMID COVID-19 Working Group. Social contacts in workplace the UK from the CoMix social contact survey. London School of Hygiene and Tropical Medicine 2021. https://cmmid.github.io/topics/covid19/reports/comix/Comix%20Report%20contacts%20in%20the%20workplace.pdf

24 Read JM, Eames KTD, Edmunds WJ. Dynamic social networks and the implications for the spread of infectious disease. J R Soc Interface 2008;5:1001–7.

25 Smieszek T. A mechanistic model of infection: why duration and intensity of contacts should be included in models of disease spread. Theor Biol Med Model 2009;6:25.

26 Danon L, Read JM, House TA, et al. Social encounter networks: characterizing Great Britain. Proc Biol Sci 2013;280:20131037.

27 Thompson HA, Mousa A, Dighe A, et al. Severe Acute Respiratory Syndrome Coronavirus 2 (SARS-CoV-2) Setting-specific Transmission Rates: A Systematic Review and Meta-analysis. Clin Infect Dis 2021;73:e754–64.

28 De Cao E, Zagheni E, Manfredi P, et al. The relative importance of frequency of contacts and duration of exposure for the spread of directly transmitted infections. Biostatistics 2014;15:470–83.

29 CDC. Masks and respirators. Centers for Disease Control and Prevention. 2022.https://www.cdc.gov/coronavirus/2019-ncov/prevent-getting-sick/types-of-masks.html (accessed 13 Jul 2022).

30 Office for National Statistics. Population estimates. 2021.https://www.ons.gov.uk/peoplepopulationandcommunity/populationandmigration/populationestimates (accessed Jan 2022).

31 Daniels S, Wei H, Han Y, et al. Risk factors associated with respiratory infectious disease-related presenteeism: a rapid review. BMC Public Health 2021;21:1955.

