## Supplementary Information for "Contact patterns of UK home delivery drivers and their use of protective measures during the COVID-19 pandemic: a cross-sectional study"

Jessica RE Bridgen; Centre for Health Informatics, Computing and Statistics, Lancaster Medical School, Lancaster University, Lancaster, UK
Hua Wei; School of Health Sciences Centre, University of Manchester, Manchester, UK
Carl Whitfield; Immunity and Respiratory Medicine, University of Manchester, Manchester, UK
Yang Han; Department of Mathematics, University of Manchester, Manchester, UK
Ian Hall; Department of Mathematics, University of Manchester, Manchester, UK
Chris P Jewell; Department of Mathematics and Statistics, Lancaster University, Lancaster, UK
Martie JA van Tongeren; Academic Health Sciences Centre, University of Manchester, Manchester, UK
Jonathan M Read; Centre for Health Informatics, Computing and Statistics, Lancaster Medical School, Lancaster University, Lancaster, UK

### Contents

***Tables*** ........................................................................................................ **2**

***Survey questions***................................................................................................ **5**

### Tables

**Table 1**: Participants' employment information. N is the number of participants who provided a response to the question.

| | **Number of participants (%)** |
|---|---|
| Current employment situation (N = 162) | |
| Self-employed and completely independent | 88 (54.3%) |
| Self-employed but receiving certain workers benefits | 14 (8.6%) |
| Employed by one company - full time | 46 (28.4%) |
| Employed by one company - part time | 11 (6.8%) |
| Employed by multiple companies | 3 (1.9%) |
| No response | 0 (0.0%) |
| Current weekly working hours (N = 162)† * | |
| 0 | 0 (0.0%) |
| 1 - 10 | 6 (3.7%) |
| 11 - 20 | 17 (10.5%) |
| 21 - 30 | 17 (10.5%) |
| 31 - 40 | 44 (27.2%) |
| 41 - 50 | 42 (25.9%) |
| 51 or more | 36 (22.2%) |
| In receipt of statutory sick leave pay (N = 151)†* | |
| Yes | 38 (25.2%) |
| No | 103 (68.2%) |
| Not sure | 10 (6.6%) |

† Question required a response from the participant.
* As a delivery driver

**Table 2:** Adjusted incidence rate ratios for number of customer contacts per shift by select variables.

| | Multivariable analysis | |
|---|---|---|
| | **Adjusted Incidence Rate Ratio (95%CI)** | **p-value** |
| Age | | |
| 18-29 | 1.65 (1.07-2.60) | 0.020 |
| 30-39 | 1.13 (0.77-1.68) | 0.525 |
| 40-49 | 1.64 (1.15-2.34) | 0.004 |
| 50-59 | 1.00 | - |
| 60-69 | 1.00 (0.60-1.75 | 0.986 |
| Sex | | |
| Female | 1.00 | - |
| Male | 0.98 (0.70-1.36) | 0.910 |
| Employment type | | |
| Self-employed and independent | 1.00 | - |
| Self-employed and receiving some benefits | 1.19 (0.74-1.97) | 0.456 |
| Employed by one company - full time | 0.66 (0.47-0.94) | 0.019 |
| Employed by one company - part time | 0.77 (0.39-1.58) | 0.404 |
| Employed by multiple companies | 0.97 (0.40-2.80) | 0.955 |
| Furthest distance from depot to delivery (miles) | | |
| 0-9 | 1.00 | - |
| 10-19 | 1.18 (0.79-1.75) | 0.422 |
| 20-99 | 1.23 (0.83-1.82) | 0.274 |
| 100-499 | 1.07 (0.60-1.91) | 0.805 |
| Weekly working hours | | |
| 1-10 | 0.47 (0.24-1.00) | 0.036 |
| 11-20 | 0.55( 0.32-0.99) | 0.018 |
| 21-30 | 0.44 (0.28-0.71) | 0.001 |
| 31-40 | 1.00 | - |
| 41-50 | 0.95 (0.63-1.43) | 0.806 |
| 51+ | 0.98 (0.65-1.48) | 0.918 |
| Items normally delivered | | |
| Small parcels only | 1.00 | - |
| Large items only | 1..24(0.55-3.13) | 0.600 |
| Small parcels and large items | 0.88(0.59-1.34) | 0.541 |
| Groceries only | 0.34(0.18-0.64) | 0.001 |

| | | |
|---|---|---|
| Takeaway only | 0.19(0.11-0.37) | <0.001 |
| Other items only | 0.43(0.24-0.80) | 0.003 |
| Other delivery type combinations | 0.58(0.38-0.93) | 0.014 |

### Survey questions

Q1a. Are you aged 18 or over?

- ◯ Yes
- ◯ No

Q1b. Do you work as a home delivery driver in the UK? (including mail, parcels, homeware, takeaway, groceries etc.)

- ◯ Yes
- ◯ No

Q1c.
**Please make sure you agree to the following before continuing with the survey:**
You currently live in the UK;
You are a home delivery driver;
You have read the Participant Information Sheet and fully understand what is expected of you within this study;
Your participation is voluntary, and you are aware that you can stop the survey at any point; You consent to Lancaster University keeping the anonymised data for a period of 10-years after the study has finished;

- ◯ **I consent to taking part in the CoCoNet: Home Delivery Driver study**

Q2. Where in the UK do you currently live?

- ◯ England
- ◯ Northern Ireland
- ◯ Scotland
- ◯ Wales
- ◯ I do not live in the UK

Q3. What is your age?

- ◯ 18 - 29 years old
- ◯ 30 - 39 years old

- ◯ 40 - 49 years old
- ◯ 50 - 59 years old
- ◯ 60 - 69 years old
- ◯ 70 - 79 years old
- ◯ Aged 80 or over

Q4. What is your sex?

The answer you give can be different from what is on your birth certificate.

- ◯ Female
- ◯ Male
- ◯ Prefer not to say

Q5. Which of the following best describes your ethnicity?

- ◯ English / Welsh / Scottish / Northern Irish / British
- ◯ Irish
- ◯ Gypsy or Irish Traveller
- ◯ Any other White background
- ◯ White and Black Caribbean
- ◯ White and Black African
- ◯ White and Asian
- ◯ Any other Mixed / Multiple ethnic background
- ◯ Indian
- ◯ Pakistani

○ Bangladeshi

○ Chinese

○ Any other Asian background

○ African

○ Caribbean

○ Any other Black / African / Caribbean background

○ Arab

○ Any other ethnic group

○ Prefer not to say

Q6. What is the first part of your home postcode?
*For example, if your home postcode was LA1 4YW then you would enter LA1.*

_______________________________

Q7. What is you highest level of education?

○ Higher education and professional/vocational equivalents

○ A levels, vocational level 3 and equivalents

○ Trade apprenticeships

○ GCSE/ O level grade A*-C, vocational level 2 and equivalents

○ Qualifications at level 1 and below

○ Other qualifications

○ No qualifications

○ Not sure

Q8. Are you self-isolating or shielding because of COVID-19?
*A vulnerable individual here refers to a clinically extremely vulnerable person.*

○ I am not self-isolating or shielding

○ Self Isolating - I have symptoms of COVID

○ Self Isolating - Someone in my household has symptoms of COVID

○ Self Isolating - Someone in my support bubble has symptoms of COVID

○ Self Isolating - Someone I have been in contact with has symptoms of COVID

○ Self Isolating - Told to do so by a contact tracer

○ Self Isolating - Told to do so by a contact tracing phone app

○ Self Isolating - In travel-related quarantine

○ Shielding - I am a vulnerable individual

○ Shielding - I live with a vulnerable individual

○ Other - *please do not include any identifying information*

______________________________________________

○ Not sure

Q9. Have you ever had to self-isolate due to COVID-19 infection (either suspected or confirmed)?

○ Yes

○ No

Q10. In which month(s) have you had to self-isolate? Tick all that apply.

- ☐ March 2020
- ☐ April 2020
- ☐ May 2020
- ☐ June 2020
- ☐ July 2020
- ☐ August 2020
- ☐ September 2020
- ☐ October 2020
- ☐ November 2020
- ☐ December 2020

- ▢ January 2021
- ▢ February 2021
- ▢ March 2021

Q11. Have you ever tested positive for COVID-19?

- ◯ Yes
- ◯ No

Q12. In which month(s) did you test positive? Tick all that apply.

- ▢ March 2020
- ▢ April 2020
- ▢ May 2020
- ▢ June 2020
- ▢ July 2020
- ▢ August 2020
- ▢ September 2020
- ▢ October 2020
- ▢ November 2020
- ▢ December 2020
- ▢ January 2021
- ▢ February 2021
- ▢ March 2021

Q13. How many other people currently live with you at home?

- ◯ 0 - I live alone
- ◯ 1
- ◯ 2
- ◯ 3
- ◯ 4
- ◯ 5 or more

Q14. How many people of each age group live with you at home?
*Do not include yourself.*

*Drop down options of 0, 1, 2, 3, 4, 5 or more for each age group.*

0 - 9 year olds
10 - 19 year olds
20 - 29 year olds
30 - 39 year olds
40 - 49 year olds
50 - 59 year olds
60 -69 year olds
70 - 79 year olds
Aged 80 or over

Q15. What do you normally deliver?

- ☐ Letters and mail
- ☐ Small parcels
- ☐ Large items (e.g. large appliances, furniture)
- ☐ Takeaway food
- ☐ Groceries
- ☐ Other - please specify ___________________________________________________

Q16. What is your current employment situation?

○ Self-employed and completely independent

○ Self-employed but receiving certain workers benefits (e.g. holiday pay, sick pay). Please specify - do not include any identifying information

___________________________________________________

○ Employed by one company - full time

○ Employed by one company - part time

○ Employed by multiple companies

Q17. How many hours do you currently work as a delivery driver per week?

○ 0 hours

○ 1 - 10 hours

○ 11 - 20 hours

◯ 21 - 30 hours

◯ 31 - 40 hours

◯ 41 - 50 hours

◯ 51 hours or more

Q18. How many deliveries did you make during your last shift?

________________

Q19. When was the last shift you worked as a delivery driver?

◯ This week

◯ Earlier this month

◯ Last month

◯ 2 months ago or more

Q20. Did you use your own vehicle to make deliveries on your last shift?

◯ Yes

◯ No

Q21. During your last shift, how many customers did you meet face-to-face?

___________________

Q22. On your last shift, which types of face-to-face contact did you make with customers? Tick all that apply.

- ▢ Brief face-to-face interaction with customer (less than 5 minutes)
- ▢ Prolonged face-to-face interaction with customer (more than 5 minutes)
- ▢ Entered a customer's property (e.g. for installation or to drop off a heavy item)
- ▢ Hand signature required from customer
- ▢ Other, please specify ______________________________________

Q23. Were you able to maintain social distance (at least 2 metres apart) from all of the customers you met on your last shift?

- Yes, all of the time
- More than half of the time
- Less than half of the time
- No, none of the time
- Not sure

Q24. During your last shift, how many people did you meet in the location where you collect items for delivery (e.g. depots or food outlets)?
*Only include those you had a face-to-face conversation with.*

______________________________

Q25. Were you able to maintain social distance (at least 2 metres apart) from everyone you met in the location where you collect items for delivery last shift?

- Yes, all of the time
- More than half of the time (2)
- Less than half of the time
- No, none of the time
- Not sure

Q26. During your last working week, what was the furthest distance you travelled from a collection point (at a depot or restaurant) to a delivery address?

- 0 - 9 miles
- 10 - 19 miles
- 20 - 99 miles
- 100 - 499 miles
- 500 miles or more

Q27. During your last working week, did you share a vehicle with a colleague to make deliveries? (e.g. for large items)

◯ Yes

◯ No

Q28. During your last working week, did you always share a vehicle with the same colleague?

◯ Yes

◯ No

Q29. What measures have you taken to prevent infection when sharing a vehicle? Tick all that apply.

- ☐ Keeping windows open
- ☐ Wearing a facemask
- ☐ Using hand sanitiser
- ☐ Other, please specify _________________

Q30. On your last shift, did you use any Personal Protective Equipment (PPE)? Tick all that apply.

- ☐ Facemask
- ☐ Gloves
- ☐ Hand sanitiser
- ☐ Other, please specify _________________

Q31. Was the PPE provided by your employers or contracting companies/platforms?

◯ Yes

◯ No

◯ Not sure

Q32. Do you think they are effective protection?

◯ Yes

◯ No

◯ Not sure

Q33. Do you think you have adequate knowledge and information about workplace COVID-19 risk?

◯ Yes

◯ No

Q34. Where do you learn about workplace COVID-19 risk? Tick all that apply.

☐ Company/platform communications
☐ Government website
☐ News and social media
☐ Other, please specify ____________________

Q35. Do you receive statutory sick leave pay for your delivery occupation?

◯ Yes

◯ No

◯ Not sure

Q36. How would you cover your living expenses if you had to take time off work due to being ill with COVID-19 or COVID-19 symptoms? Tick all that apply.

☐ Special COVID-19 funding scheme provided by the contracting companies/platforms
☐ Government self-employment income support scheme
☐ Social benefits
☐ Other, please specify ___________________

Q37. Since March 2020, have you worked whilst being ill with COVID-19 symptoms, or with a member of your household having a suspected or confirmed case of COVID-19?
*COVID-19 symptoms include: a high temperature, a new continuous cough, a loss or change to your sense of smell or taste*

◯ Yes

◯ No

◯ Not sure

Q38. What was the main reason you still went to work? Tick all that apply.

☐ Financial reasons
☐ Unable to find someone to cover my shifts
☐ Asked to or encouraged to carry on working by my employers
☐ Other, please specify ________________________________
